# MULTIPLE PRIMARY CANCERS IN LUNG CANCER PATIENTS IN ATLANTIC CANADA AND IMPLICATIONS FOR SCREENING AND MANAGEMENT

**DOI:** 10.1101/2025.01.07.25320152

**Authors:** Kassandra M. Coyle, Yingtong Gao, Rowan E. Murphy, Victor Martinez, Aaron Goodarzi, Graham Dellaire, Alison Wallace

## Abstract

**Background:** Lung cancer incidence rates in Atlantic Canada significantly exceed those of other Canadian provinces, with underlying causes remaining poorly understood. This regional disparity suggests potential genetic or environmental factors unique to Atlantic Canada. Here we present for the first-time data indicating that accompanying the high lung cancer rates in Atlantic Canada is an intricate landscape of multiple malignancies. This represents a phenomenon of multiple primary cancers that is unprecedented in the literature and presents unique challenges in the diagnosis and treatment of individuals diagnosed with lung cancer in Atlantic Canada.

**Methods:** We performed a retrospective chart review of 1,151 patients referred to the thoracic surgeons at the Queen Elizabeth II Health Sciences Centre between 2019 and 2023 with the new diagnosis of lung cancer. The primary focus was to assess the incidence of multiple primary cancers (as documented in pathology reports) and analyze demographic, clinical, and geographical factors through Fisher’s exact tests and Student’s t-tests.

**Results:** Forty-three precent (43.3%) of Atlantic Canadian patients presented with multiple primary cancers. Of 1,949 cancers identified, 1,440 were of primary lung origin. Sixty percent (60.0%) of our cohort was female. Thirty-one percent (31.3%) were self-reported current tobacco smokers, 50.3% were former tobacco smokers, and 18.4% reported no tobacco use. The average age and body mass index (BMI) at first diagnosis was similar between those patients with and without multiple primary cancers, averaging 67 years and 27.7 kg/m^2^, respectively. Thirty-nine percent (39.2%) of our study participants lived in more densely populated centers (>100,000 people), 10.2% in intermediate centers (∼98,000 people) and 50.7% in more sparsely populated centers (<55,000 people).

**Conclusions:** We have uncovered an unprecedented phenomenon in Atlantic Canada of a cohort of patients with lung cancer having multiple primary cancers of different tissue origin at a rate that is four times greater than previously described. The implications of this study are that Atlantic Canadians are disproportionately burdened with cancers of multiple origins that are not explained by smoking rates or BMI. Future studies will investigate the contribution of genetic predisposition, environmental exposures, and socioeconomic factors unique to Atlantic Canada underlying the high rates of multiple primary cancers documented here in a lung cancer cohort. Such studies will be essential to develop effective cancer prevention and screening strategies in Atlantic Canada.

## INTRODUCTION

Lung cancer remains a critical public health challenge in Canada, representing the most frequently diagnosed cancer and the leading cause of cancer-related mortality^1^. Particularly noteworthy are the elevated lung cancer age-standardized incidence rates within Atlantic Canada, where figures for 2023 are anticipated to range between 64.4-80.2 cases per 100,000 individuals, in contrast to 58.4-60.1 per 100,000 in other Canadian provinces^1^. The etiology behind these heightened rates in Atlantic Canada remains elusive.

Familial clustering of predisposing disease gene alleles occurs in Atlantic Canada, possibly due to a founder effect, including genes linked to cancer predisposition^2–4^. However, these genetic factors, while significant, cannot fully account for the elevated cancer incidence across various cancer types in this region. Tobacco smoking and obesity are known contributors to cancer incidence; however, despite similar rates of smoking and obesity, the Atlantic Region has a higher lung cancer rate than Quebec^4^. Finally, the unique geography of Atlantic Canada may also expose its population to carcinogenic agents like radon and arsenic through air and water (respectively), exposures that are known to increase occurrences of lung cancer and other solid tumors^5–7^. Likely reflecting a unique interplay of genetic, environmental, and socioeconomic factors, we have identified a large cohort of patients with multiple primary cancers of different tissue origin who at some stage of their journey with multiple cancers are diagnosed with lung cancer and referred to a thoracic surgeon.

We suggest this phenomenon of multiple primary cancers, without a clear genetic origin, over the lifetime of this cohort of Atlantic Canadians represents a previously undescribed “Atlantic Cancer Syndrome”. Thus, the intersection of augmented cancer rates in Atlantic Canada and the potential escalated risk of exposure to IARC group 1 carcinogens like radon and arsenic prompts future inquiry into whether an amalgamation of genetic susceptibility, environmental elements and/or socioeconomic factors underlies the region’s cancer statistics (Figure 1).

**Figure 1.**
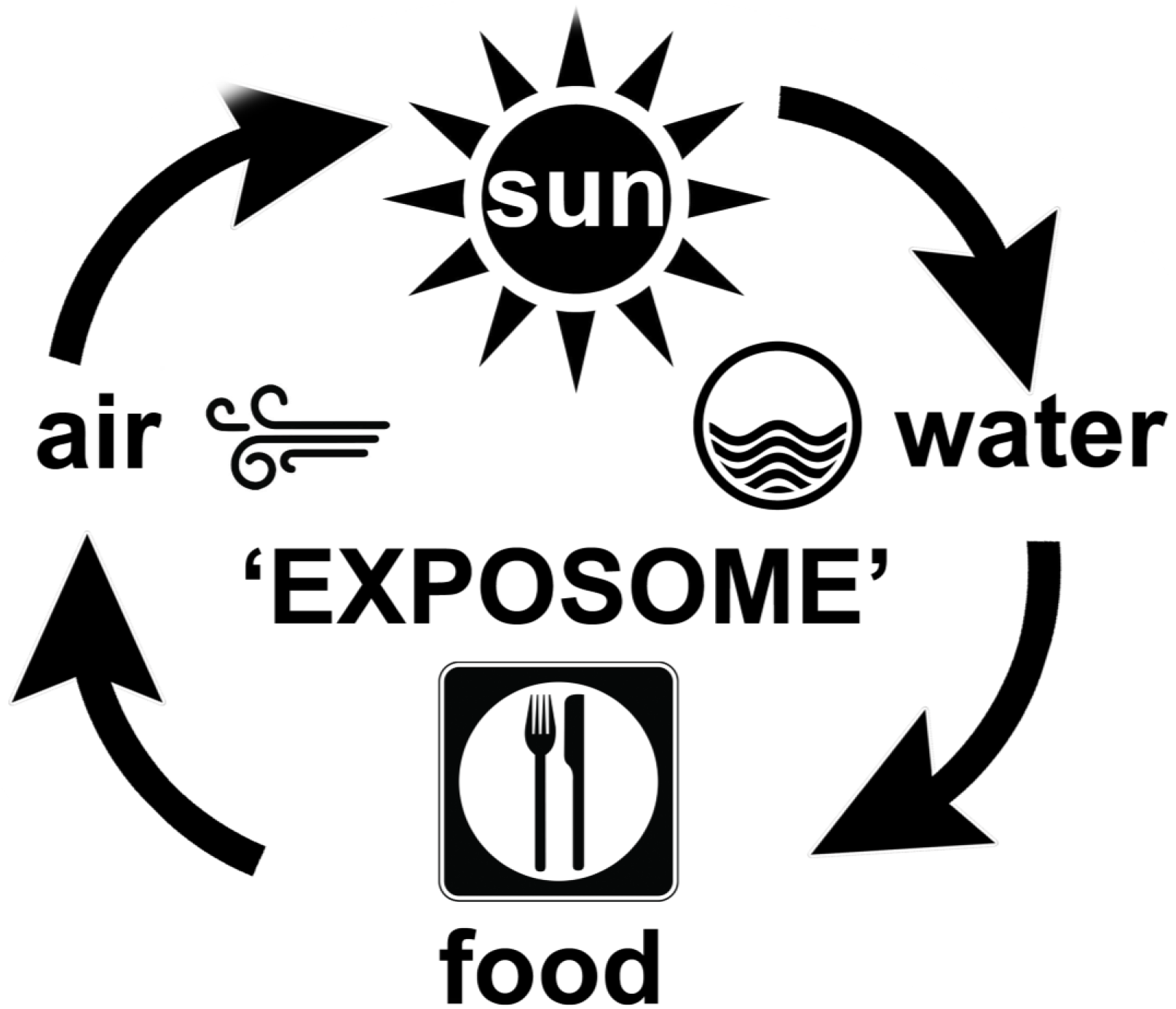
The Exposome - Something in the air and water: Studying the cancer predisposing genetic and environmental factors that underlie the high lung cancer rates in Atlantic Canada and the emergence of Atlantic Cancer Syndrome.

## METHODS

All lung cancer referrals in Nova Scotia are triaged to the thoracic surgeons at the Queen Elizabeth II Health Sciences Centre (QEII HSC) in Halifax, which also serves as a referral center for patients from Prince Edward Island and New Brunswick. Study participants were retrospectively identified from this referral population between January 1, 2019, and December 31, 2022. The classification of “primary cancer” was based on the type and stage of cancer described in tumor biopsy or tumor resection pathology reports. Primary cancer was defined as cancerous cells originating from the tissue of interest (e.g., adenocarcinoma of the left lower lobe in a lung tumor biopsy), whereas metastatic cancer was identified when cells originated from tissue outside the lung (e.g., metastatic adenocarcinoma of colorectal origin in a lung tumor biopsy). Data on date of birth, sex, tobacco smoking status, BMI, and postal forward sortation area were collected for subsequent analysis. Ethics approval for this study was obtained from the Nova Scotia Health Research Ethics Board.

Statistical analysis was performed using R Statistical Software version 4.4.2. Continuous variables (e.g., age and BMI) are expressed as medians with ranges, while categorical variables (e.g., sex and smoking status) are expressed as percentages. The Fisher’s exact test was used for the comparison of categorical variables, and the Student’s t-test was applied for continuous variables. A P-value of <0.05 was considered statistically significant. We used the Benjamini-Hochberg procedure to control for multiple comparisons.

## RESULTS

Chart reviews of 1,151 patients treated by the thoracic surgeons serving Atlantic Canada revealed that 43.3% had multiple (>2) primary cancers over their lifetime, including both synchronous and metachronous cancers (Figure 2A). Specifically, 29.2% had double primaries, 9.6% had triple primaries, and 3.2% had quadruple primaries. Additionally, some patients had more than six primary cancers throughout their lifetime.

**Figure 2.**
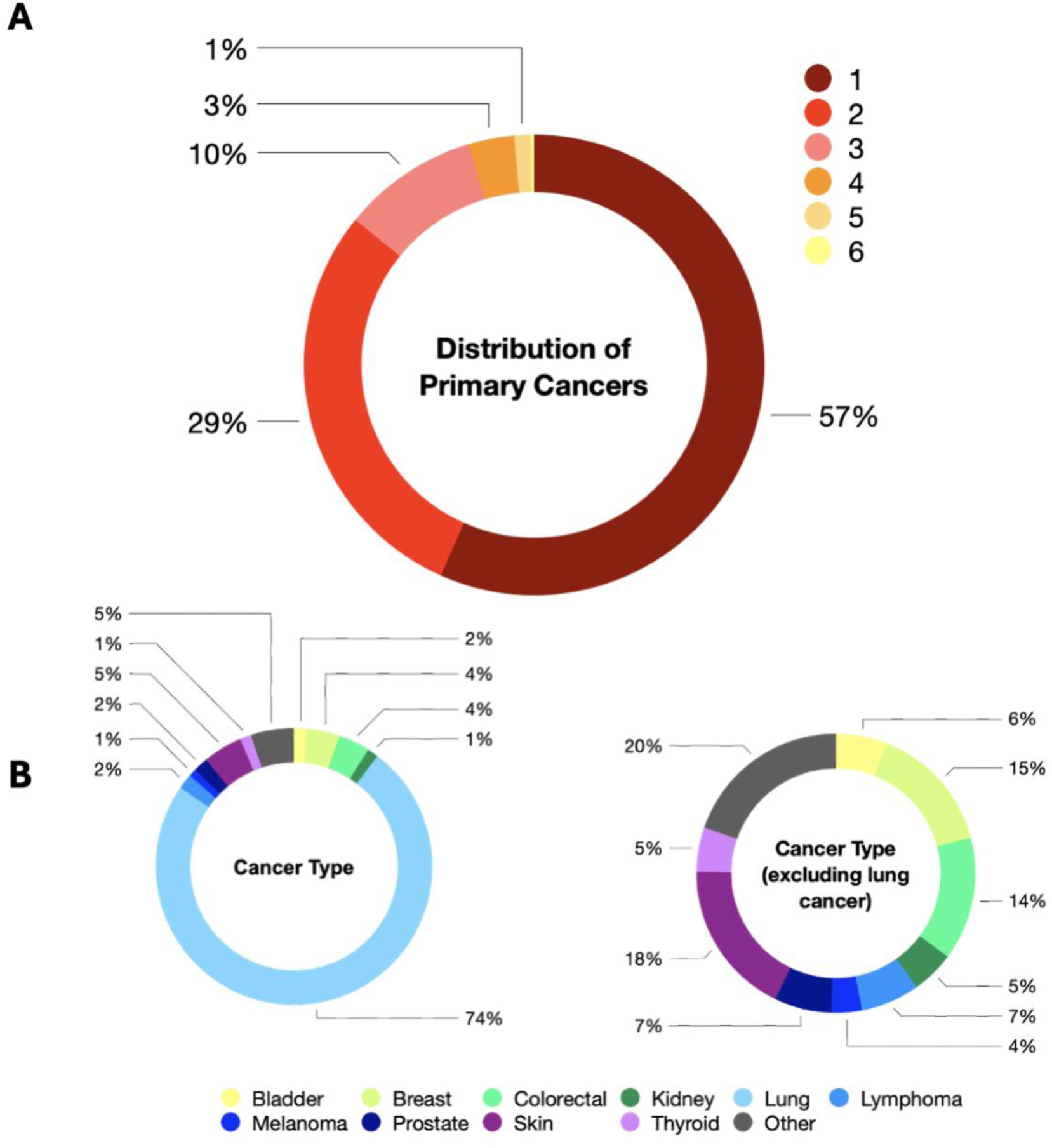
Cancer Distribution. A. Distribution of primary cancers in the study cohort. B. Distribution of cancer type in the study cohort.

A total of 1,949 cancers were identified in our patient cohort, with 1,440 for lung origin. Other than lung cancer, non-melanoma skin cancer (351), breast cancer (292), colorectal cancer (274) and prostate cancer (136) were found to be most prevalent (Figure 2B). This is similar to the Canadian Cancer Registry projections for 2024 where lung cancer is expected to be most the prevalent diagnosis at 13% followed by breast (12.5%), prostate (11.3%) and colorectal (10.2%)^8^. Simple comparison of percentages from our study to the Canadian Cancer Registry is subject to potential bias from the use of lung cancer as an inclusion criterion for our study and was thus omitted.

The distribution of female and male patients was statistically significant (p=0.039). Six hundred and ninety-two (60.0%) study patients were female compared to 461 (40.0%) males. This distribution between female and male patients was consistent between all subgroups in our study (Table 1). Three hundred and sixty (31.7%) patients were self-reported current tobacco smokers at the time of their first diagnosis, 579 (49.8%) were former tobacco smokers and 214 (18.4%) patients indicated they had never smoked tobacco (Table 1). Former smokers had significantly more multiple primaries than either never smokers or current smokers (p=0.006). This relationship between tobacco smoking status and number of primaries was consistent across all sub-groups within our study. There were no differences in age at first diagnosis or BMI at first diagnosis across all study groups (Table 1).

**Table 1.** Demographic characteristics of the study cohort.

|  | **1 Primary** | **2 Primaries** | **3 Primaries** | **4 Primaries** | **5+ Primaries** |
| --- | --- | --- | --- | --- | --- |
| **Distribution of primary cancers – no (%)** | 652 (56.7%) | 336 (29.2%) | 110 (9.6%) | 37 (3.2%) | 16 (1.4%) |
| **Sex – no (%)** |  |  |  |  |  |
| **Female** | 375 (54.2%) | 203 (29.3%) | 81 (11.7%) | 24 (3.5%) | 9 (1.3%) |
| **Male** | 277 (60.3%) | 133 (29.0%) | 29 (6.3%) | 13 (2.8%) | 7 (1.5%) |
| **Smoking status – no (%)** |  |  |  |  |  |
| **Never smoker** | 134 (62.6%) | 50 (23.4%) | 16 (74.8%) | 9 (4.2%) | 5 (2.3%) |
| **Former smoker** | 301 (52.0%) | 187 (32.3%) | 63 (10.9%) | 23 (4.0%) | 5 (0.9%) |
| **Current smoker** | 219 (73.2%) | 50 (16.7%) | 16 (5.4%) | 9 (3.0%) | 5 (1.7%) |
| **Age at first cancer diagnosis – Median (range)** | 68 (18-88) | 69 (35-85) | 68 (39-82) | 72 (34-80) | 72 (24-75) |
| **BMI at first cancer diagnosis – Median (range)** | 27.1 (15.1-59.2) | 25.4 (15.8-49.5) | 26.4 (15.2-51.9) | 28.7 (20.6-48.0) | 27.8 (21.9-43.9) |

To help integrate residential status in our study considerations, patients within Nova Scotia were mapped based on postal forward sortation area provided in the patient chart. 39.2% of our study participants lived in Halifax County (population = 460,232 people) and 10.2% lived in Cape Breton County (population = 98,635 people), the two largest urban cities within the province^9^ (Figure 3A). The remaining 50.7% of our study participants were spread across the other 16 counties, all with a population less than 55,000 people^9^ (Figure 3A). We also compared our postal forward sortation area distribution to the four Nova Scotia Health Zones (Figure 3B). 43.5% of study patients were within the central health zone, which was a significantly greater proportion of patients than all other zones (p= 0.013, p=0.001, p=0.002; Figure 3B and Figure 3C).

**Figure 3.**
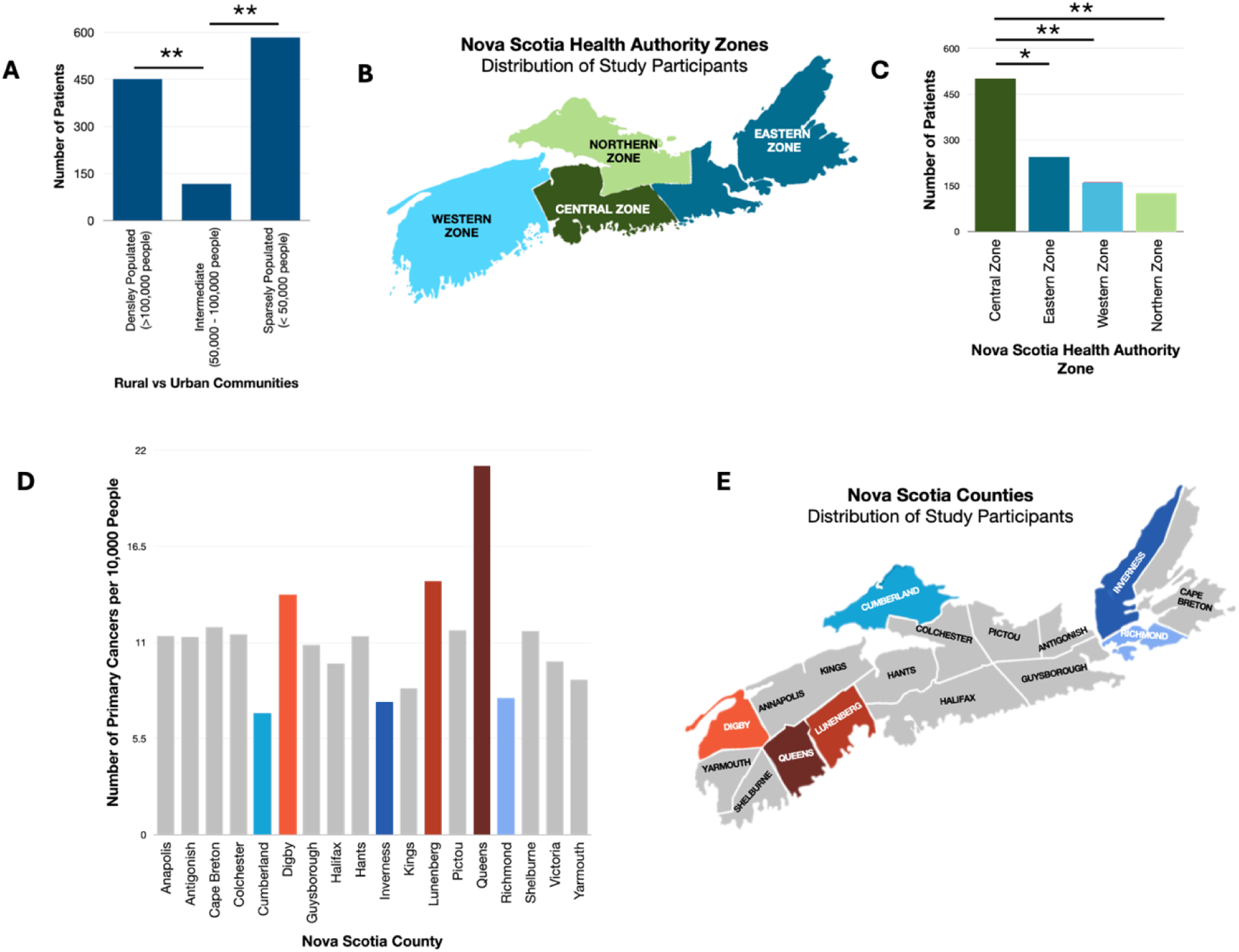
Population distribution of cohort. A. Density of study participants grouped into rural (<55,000) semi-urban (55,000-100,000) and urban (>100,000) communities. B. Illustration of Nova Scotia Health Authority Zones^9^. Summary of patient distribution by healthcare zone (C) and county (D, E) based on postal forward sortation area.

Due to the differences in number of study participants compared to population distribution, the population was normalized for 10,000 individuals, based on the 2021 Government Nova Scotia Census Report^9^. Study participants were grouped by county, after being normalized for 10,000 individuals (Figure 3D). Queens County had the highest prevalence of patients with multiple primary cancers, at 21.1 per 10,000 individuals, followed by Lunenburg County at 14.5 and Digby County at 13.7. In contrast, Cumberland County (6.91), Inverness County (7.45), and Richmond County (7.69) had the lowest prevalence (Figure 3D, E).

## DISCUSSION

We have identified that during the study period 43.3% of patients with lung cancer referred to the thoracic surgeons serving the Atlantic Provinces have multiple primary cancers of different tissue origin (Figure 2A). These numbers are staggering compared to previously reported literature. Ge et al., describe double primary cancers occurring in 11% of patients, triple primaries in <0.5% and quadruple primary cancers in <0.1%^10^. Similarly, Jayarajah et al., report that double primary cancers occur in 16% of patients and quadruple in <0.1%^11^. Therefore, our study describes a prevalence of multiple primary cancers in Atlantic Canada that is almost four times greater than what has been previously stated in the literature; a phenomenon we are terming Atlantic Cancer Syndrome due to the uncertain etiology of clustered primary cancers in our cohort. It should be noted that we first became aware of this phenomenon after employing positron emission tomography for evaluation of lung cancer patients during initial work-up, which enabled the facile identification of multiple synchronous primaries at first diagnosis. Thus, our data set is enriched for patients with synchronous primary cancers at diagnosis, as compared to previous studies of mostly asynchronous primary cancers in different tissues, making our findings all the more astonishing. As such, our findings indicate an urgent and unmet need to identify the etiology of the Atlantic Cancer Syndrome phenomenon we are reporting here.

Genetic predisposition could be one factor, given the region’s limited genetic diversity, which may have resulted in the enrichment of deleterious genetic sequences and founder effects that increase susceptibility to certain cancers. We also observed a significant enrichment of smokers among those with multiple cancers. In particular, the most common synchronous primary cancer associated with lung cancer in our cohort was non-melanoma skin cancer, and multiple studies have indicated an increased risk of non-melanoma squamous cell skin cancer among smokers^12,13^. In our study, we have considered that former smokers may be more likely to engage in preventive health behaviors such as seeking medical care and undergo routine cancer screening. This increased surveillance could have led to earlier detection of multiple primary cancers in these individuals, which may partially explain the observed discrepancy. Additionally, former smokers tend to survive longer than current smokers, providing a longer time window for new primary cancers to emerge and be detected. Of note, the Quebec region of Canada is also subject to reduced genetic diversity and high rates of smoking similar to Nova Scotia and other Atlantic Provinces^14^. Yet, similar reports have not emerged regarding high levels of synchronous multiple primary cancers in the Quebec population. Therefore, we are left to assume that the high cancer rates observed in Atlantic Canada cannot be solely attributed to smoking or genetic factors. Thus, we propose there may be an interplay between smoking, genetic predisposition and environmental exposures in the region; exposures that are critical to our understanding of the etiology of the elevated cancer rates in the Atlantic region.

Due to its geology, people living in Atlantic Canada are exposed to carcinogens in the air they breathe and the water they drink – radon, arsenic and air pollution are environmental risk factors of interest that are known to be elevated within the Atlantic region^5–7^. We saw a clustering of study participants in Queens County, Digby County and Lunenburg County, leading us to question whether environmental factors in these regions are putting individuals at greater risk of cancer. Further, our data shows that the entirety of Atlantic Canada has an increased prevalence of multiple primary cancers when compared to previously reported literature,^10,11^ which reduces the probability of genetic founder effects being the major contributor to the observed malignancy rates. Again, this raises the question of whether environmental factors, rather than any genetic predisposition alone, are contributing to heightened cancer rates within Atlantic Canada.

Radioactive radon gas is the second leading cause of lung cancer for Canadians; although minimally dangerous at the low levels found in outside air, indoor environments can often contain high levels of radon in modern buildings that have increasingly retained and concentrated this gas^15–19^. Long-term exposure to radon not only increases lung cancer risk but also intensifies the impact of tobacco use on lung cancer risk by a factor of 17, amplifying the negative health consequences of tobacco smoking^,18,20–23^. Radon exposure and its impact on cancer susceptibility varies across populations based on age, sex, region, occupation, socioeconomics, and genetics. Nonetheless, Canadians are among the 2^nd^ overall most radon-exposed people worldwide, and exposure in residential areas continues to rise^17^. In fact, Prairie Canada, Northern Canada, and Atlantic Canada have the second, third, and fourth highest residential radon levels in the world, respectively^24^. In addition, people living in rural communities experience 31.2% greater average residential radon levels relative to urban equivalents, which is thought to be attributed to the increased use of groundwater wells in these communities^25,26^. Postal forward sortation area data in this study revealed that 60.82% of our patients lived in a community with a population <100,000, suggesting a potential risk to radon exposure for patients in these communities (Figure 3A).

The government of Nova Scotia has published a report on the risk of indoor radon exposure in homes^27^. More recently, the 2024 Cross-Canada Survey of Radon Exposure in Residential Buildings^28^ was published which further highlights significant radon risks in Atlantic Canada, with approximately 1 in 3 (33.3%) households in the region having radon levels at or above the Canadian guideline of 200 Bq/m³. The average radon level in Atlantic Canadian homes is 116.8 Bq/m³, the second-highest among all Canadian regions. Nova Scotia, in particular, stands out, with 36.8% of residential properties exceeding the 200 Bq/m³ threshold, and an average radon level of 125.3 Bq/m³. The data from this report clearly show that all regions of the province have some level of risk to indoor radon exposure. However, the city of Halifax and parts of southwestern Nova Scotia are the highest risk regions, and recently a distinct bias has been shown for higher residential radon levels in rural homes compared to urban, potentially due to the proximity of water wells that provide a conduit for radon in soil gases^29^. Interestingly, Queens County, Lunenburg County and Digby County are all located within southwestern Nova Scotia and shown to have the greatest proportion of multiple primaries in our study (Figure 3D, E). One of the next steps for this project is to quantify radon exposure in the study cohort to explore the relationship between radon exposure and the incidence of multiple primary cancers at an individual level.

Like radon, arsenic exposure can vary considerably by region. Twelve percent of Atlantic Canada’s private water wells exceed Canadian maximum allowable arsenic concentration in drinking water (≥10 μg/L), and excess arsenic body burden has been verified in Atlantic Canadians, with disproportionately high arsenic-linked cancer rates in the region^30–32^. A previous study reported that Nova Scotians have an increased risk of developing bladder and kidney cancers as a result of exposure to drinking water contaminated with arsenic, even in areas found to be within the current maximum acceptable concentration (<10 μg/L)^33^. Similarly to radon exposure, Nova Scotia’s southwest region (which overlaps with Queens County, Lunenburg County and Digby County) was found to have the highest relative risk (Figure 3D, E)^33^. Other high arsenic areas include parts of the Yukon, British Columbia, Nunavut, and Southern Ontario^34^. Arsenic-related lung tumors have been described as a molecularly distinct entity^35,36^. This molecular signature is particularly evident in lung squamous cell carcinomas among never-smokers, which is a rare combination due to the high prevalence of smokers among patients displaying this particular histological subtype.

Atlantic Canada is also known to have increased levels of air pollution, specifically PM2.5 which refers to air-borne particles or droplets that are 2.5 microns in size^37^. Air pollution problems are global and increasing: the World Health Organization estimates that 99% of all people worldwide currently reside in areas that exceed acceptable air quality limits^38^. Woodsmoke and traffic-related fuel combustion are the two biggest contributors to air pollution in Canada^39–41^. These processes generate complex mixtures of particulate matter of variable size (e.g., 2.5-10 microns) that are harmful to lung health and potent lung carcinogens^42–46^. In fact, lung cancer risk in never-smokers increases directly with PM2.5 exposure^47^. Never-smokers accounted for 18.56% of our study group, with 80 (37% of all never-smokers) of these individuals having multiple primaries (Table 1).

In addition to environmental exposures, socioeconomic factors could predispose Atlantic Canadians to an increased risk of cancer development. The 2016 census data released by Statistics Canada revealed that Nova Scotia and New Brunswick had the highest rates of low-income families (22%), in the country^48^. One study revealed that Nova Scotia had over 32,000 individuals living in areas with extreme levels of poverty and deprivation resulting in 14,693 premature deaths over an 11-year period^49^. Relevant to this study, it has been reported that the risk of developing lung cancer is 50-80% higher for patients living in low socioeconomic status^50^. There is evidence of multiple inequalities relating to diagnosis, treatment received, and survival with several studies reporting socioeconomic barriers associated with lung cancer screening ultimately leading to delayed diagnosis and worse patient outcomes^51–53^. This link between lower socioeconomic status and increased risk of lung cancer has been validated from European and American studies. Although smoking status was not associated with increased risk of multiple primary cancers in our study, a report on lung cancer and equity in Canada determined that people with lower income and people living in rural and remote communities are more likely to smoke tobacco, which is associated with a higher incidence of lung cancer^49^. Further, people with lower income are more likely to be diagnosed later in their disease, leading to worse outcomes, and even if diagnosed at an earlier stage are less likely to receive curative surgery^54^. Due to the retrospective nature of our study, we were unable to access socioeconomic information from our participants. We believe there is merit for a future study to investigate the relationship between multiple primaries, cancer outcomes and socioeconomic factors (such as household income, immigration status, race and literacy level).

Uniquely, Nova Scotia’s healthcare system is divided into four distinct healthcare zones, which aim to coordinate and deliver healthcare services to their population. Significant variability in access, infrastructure, resources, and expertise exists in the various healthcare zones, all of which have the potential to impact early detection of lung cancer^50^. Variations in treatment also exist within the healthcare zones, with the only tertiary care center and the entire thoracic surgery service for Atlantic Canada located in the Central Zone^55^. Geographic factors can influence individual ease in accessing healthcare services, as rural and remote communities often face transportation challenges which may lead to delays in seeking medical attention or accessing treatment^56^. Our study revealed that 56.5% of our study patients were based outside of the predominately urban Central Zone in Nova Scotia. Another limitation to our study is that we did not collect patient outcomes, however, a future study will include patient outcome in relation to their Nova Scotia Health Zone.

In our study, we also found that former smokers experience a higher incidence of multiple primary cancers compared to current smokers. This finding is a particular point of interest for the design and implementation of prevention strategies, as simple cessation of smoking may not be enough to fully mitigate cancer risk^57^. In future, this apparent paradox may be better understood through the lens of persistent inflammation and biological changes induced by smoking, as well as behavioral factors.

Research on chronic obstructive pulmonary disease highlights a critical phenomenon: the inflammatory response triggered by cigarette smoke exposure does not resolve immediately after cessation^58,59^. In fact, this inflammation can persist for up to 10 years or longer, depending on the duration and intensity of smoking exposure. Chronic inflammation is a well-established driver of carcinogenesis. It creates a pro-tumorigenic microenvironment characterized by oxidative stress, DNA damage, and the release of cytokines and growth factors that promote cell proliferation, angiogenesis, and genomic instability. These processes increase the likelihood of developing new primary cancers in individuals who have stopped smoking, particularly in tissues already damaged by years of exposure.

This persistent inflammatory state may be further exacerbated by environmental exposures such as radon, arsenic, and particulate matter (PM2.5); all highly prevalent in Atlantic Canada. These carcinogens can interact with the pre-existing inflammatory and damaged tissue environment, compounding the risk of carcinogenesis; exemplified by the recent discovery of inflammatory cascades initiated by PM2.5 exposure that promote lung cancer^57^. Environmental risk factors thus play a synergistic role, amplifying the impact of smoking-related tissue injury and inflammation on cancer risk.

## CONCLUSION

Our study reveals an unprecedented rate of multiple primary cancers in Atlantic Canadian lung cancer patients, with 43.3% of cases presenting multiple malignancies, a rate four times higher than previously reported. Of 31,000 new diagnoses across Canada in 2023, 4,000 were in Atlantic Canada and mostly at advanced stages, by which five-year survival rates are less than 20%. This abysmal statistic highlights the urgent need for prevention strategies and screening programs to detect lung cancer early.

Currently, a person will only be screened for lung cancer in Nova Scotia if they are between 50 and 74 years old and have a significant smoking history. But factors other than smoking now account for one-fifth to one-third of new lung cancer cases. The “Atlantic Cancer Syndrome” phenomenon uncovered here, is only the start of our team’s investigation into cancer discrepancies in the Atlantic Region compared to other provinces in Canada. The bias of enrichment of cases in rural areas of Nova Scotia, which are also burdened by higher exposure to radon^26^ and arsenic^60^ strongly indicates that environmental exposures may provide some clues as to the etiology of multiple primary cancers in Atlantic Canada. Furthermore, they argue for the future importance of measuring environmental exposures as a component of improved screening criteria for lung cancer, particularly in never-smokers.

To address this next challenge, our research team is developing personalized risk scores based on estimated lifetime exposures to radon, arsenic, and fine particulate matter (PM2.5). We are carefully defining the study population and collecting biological samples to create innovative tests that measure cumulative exposure to these environmental carcinogens. By analyzing tissues and toenails, we aim to uncover biomarker evidence of exposure, refining screening criteria to better identify individuals at risk of lung cancer. Ultimately, we envision environmental “exposome” risk scores as a transformative tool for prioritizing Canadians at the highest risk for lung cancer, enabling prevention strategies and earlier, more targeted screening interventions.

## Data Availability

All data produced in the present study are available upon reasonable request to the authors.

